# Knowledge and Impact of COVID-19 on Middle-Aged and Older People living with HIV in Lima, Peru

**DOI:** 10.1101/2021.04.23.21255998

**Authors:** Monica M. Diaz, Diego M. Cabrera, Marcela Gil-Zacarías, Valeria Ramírez, Manuel Saavedra, Cesar Cárcamo, Evelyn Hsieh, Patricia J. García

## Abstract

COVID-19 has had an unprecedented worldwide impact, and Peru has had one of the highest COVID-19 case rates despite implementation of an early strict nationwide quarantine. Repercussions on Peru’s healthcare system may impact vulnerable populations, particularly people with HIV (PWH). We explored the knowledge of COVID-19 and the socioeconomic and health impact of the pandemic among middle-aged and older PWH. A cross-sectional telephone survey was administered to 156 PWH age ≥40 years receiving care in one of two large HIV clinics in Lima, Peru. The majority of PWH (age 52±7.7 years, 41% female, 65% completed secondary school or less) were knowledgeable regarding COVID-19 symptoms and prevention methods. Nearly half of those employed prior to the pandemic reported job loss. Female sex (unadjusted prevalence ratio [PR] 1.85 [95%CI 1.27-2.69]), low educational level (PR 1.62 [1.06-2.48]) and informal work (PR 1.58 [1.06-2.36]) were risk factors for unemployment but not in adjusted models. Increased anxiety was reported in 64% and stress in 77%. COVID-19 has had a substantial socioeconomic and mental health impact on PWH living in Lima, Peru, particularly those with lower educational levels and informal workers. Efforts are needed to ensure continued medical care and socioeconomic support of PWH in Peru.

## BACKGROUND

The Coronavirus Disease 2019 (COVID-19) pandemic has had an unprecedented worldwide impact with significant repercussions in the Latin American region. Although Peru ranks 43^rd^ in the world by population size,^1^ it has one of the highest COVID-19 case counts with over 1.7 million reported cases as of April 2021.^2^ The Peruvian government declared a national state of emergency on March 15th, 2020, with a nationwide lockdown implementing strict control over the population’s ability to go outdoors (except to purchase food or pick up medications), a nightly curfew, and closure of national borders.^3^ This quarantine helped flatten the curve of infections in the country, but also had a significant impact on non-COVID-19 healthcare availability.^4^

The COVID-19 pandemic has made each step of HIV care challenging, risking the improvements achieved in HIV-related outcomes over the past decade in Peru.^5^ Detection of new cases of HIV has increased largely due to widespread implementation of HIV screening programs in Peru.^6^ However, during the first half of 2020, only 927 new cases of HIV were diagnosed, a drastic decrease compared with an incidence of 8,105 new HIV diagnoses reported the year prior, likely reflecting a decline in HIV testing among those at-risk for HIV during the pandemic.^6^

The pandemic has also had a substantial socioeconomic impact on the general population in Peru, particularly among informal workers who make up nearly 70% of the Peruvian workforce, and who are burdened with unstable incomes and inadequate healthcare coverage.^7^ From May through July 2020, the Peruvian National Institute of Statistics and Informatics (INEI) reported 6 million people had lost their employment.^8^ It is essential to determine the socioeconomic impact that COVID-19 has had on particularly vulnerable populations, such as those who are informally employed and those with lower educational levels, especially among PWH who are known to be at higher risk for medical comorbidities and poor mental health outcomes.^9,10^ Very few studies from the Latin American and Caribbean (LAC) region^11,12^ address the socioeconomic and health impact of the pandemic, or the knowledge of and attitudes towards COVID-19 infection among PWH. We present findings from a study evaluating knowledge regarding COVID-19, and the socioeconomic and health impacts of the pandemic among middle-aged and older PWH from Lima, Peru.

## MATERIAL AND METHODS

### Study design and population

We conducted a cross-sectional study between July and August 2020 among PWH receiving routine healthcare at one of two large HIV clinics located in Lima, Peru. All participants previously enrolled in one of two cross-sectional studies (on aging-related musculoskeletal and neurocognitive comorbidities among PWH) were invited to participate. There were no exclusion criteria other than those of the parent studies (age <40 years, nationality other than Peruvian nationality). Participants were re-contacted by telephone and invited to participate in the telephone survey.

### Ethics

This study was reviewed and approved by the ethics committees of the Yale School of Medicine and Universidad Peruana Cayetano Heredia. All participants provided verbal informed consent after a comprehensive explanation of the procedures by phone prior to study enrollment.

### Sources of Data

Our survey consisted of 34 questions, divided in four sections, adapted from other validated survey studies^13–15^: (a) knowledge and perceptions of COVID-19 infection, (b) socioeconomic and health impact of the COVID-19 pandemic, (c) COVID-19 infection history and diagnosis, and (d) occupation and employment history (Supplementary Table I). Questions were translated from English into Spanish by two fully bilingual investigators (MMD, DMC), and was corroborated by two native Spanish-speaker study team members (VR, MS). For all patients, demographic information (age, sex, educational level, district of residence) was obtained from their prior project records with their consent.

### Data analysis

Means and standard deviations were used to report descriptive statistics of continuous variables and frequencies and percentages for categorical variables. Univariate and multivariable (adjusted for relevant covariates, such as sex, educational level and/or occupation type) were performed using generalized linear model with family Poisson and link log to calculate prevalence ratios (PR) and their corresponding 95% confidence intervals (95% CI). All variables that were significant in the univariate analyses were included as covariates in the multivariable analyses. Statistical analyses were performed using STATA (College Station, TX, USA) and JMP Pro version 14.2.0 (Cary, NC, USA).

## RESULTS

### Sociodemographic and clinical characteristics

A total of 156 consented to participate in this study (age 52±7.7 years, 41% female) (Table 1). Eleven percent completed up to primary school, 54% more than primary school but up to secondary school, and 35% completed above secondary school. Occupations also significantly differed between sexes. The most common occupation among women was being a housewife (43.8%). We found that around 44% of the cohort was formally employed, and the rest had informal work (26.6%) or were unemployed prior to the pandemic (29.2%); Table 1.

**Table 1.**
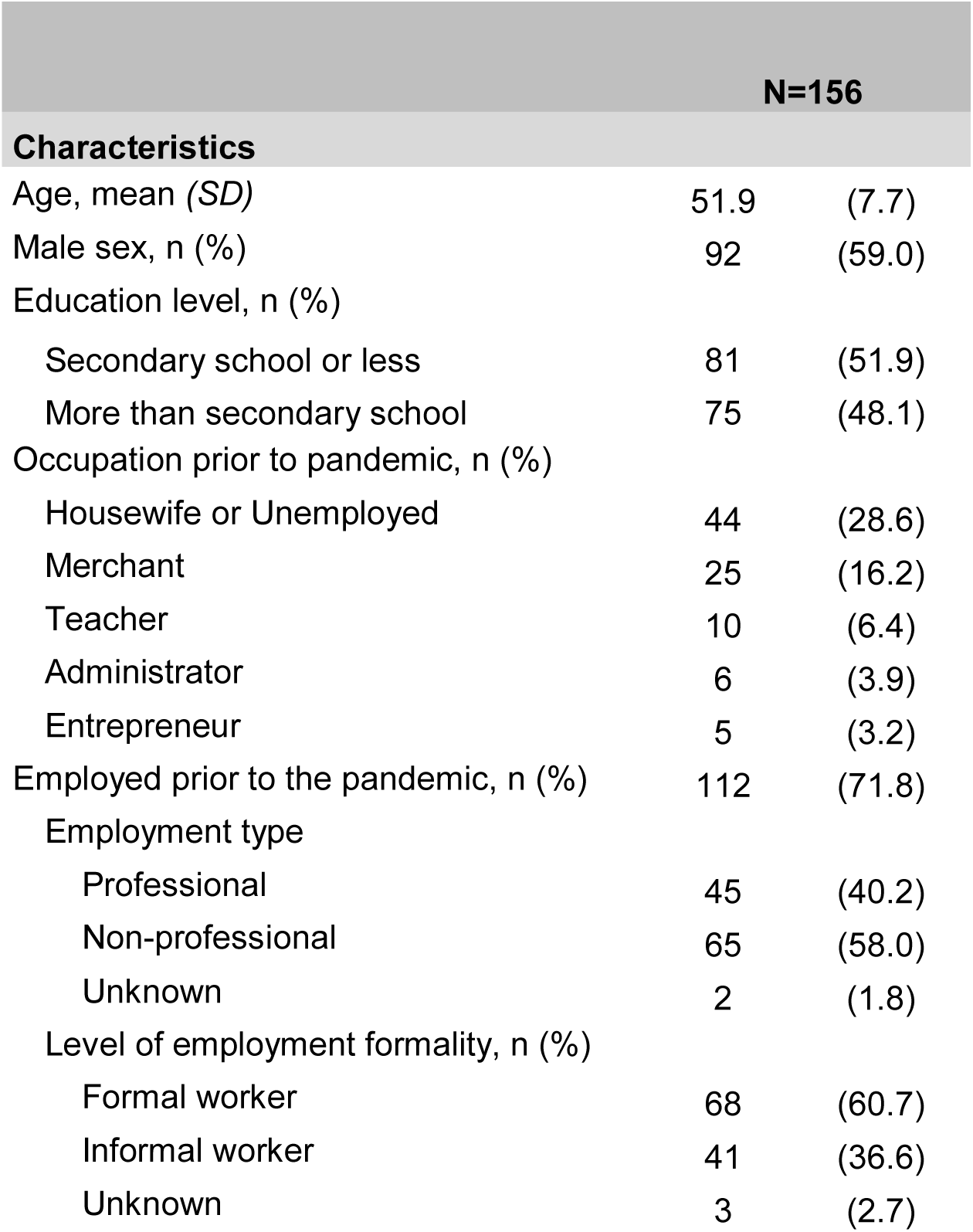
Sociodemographic characteristics of participants (N=156)

More than 10% of patients reported symptoms of COVID-19, but only four participants reported testing positive for COVID-19 infection, and no one with self-reported infection reported hospitalization (Table 2). No differences in COVID-19 infection rates were observed by educational level (p=0.746) or employment type (formal vs informal work, p=0.824).

**Table 2:** Socioeconomic and health impact of the COVID-19 pandemic on middle-aged and older PWH living in Lima, Peru by Educational Level^a^.

|  | Secondary school or less<br>n (% Yes)<br>n=81 | Above secondary school<br>n (% Yes)<br>n=75 | Prevalence Ratio<br>(95%CI)<sup>a</sup> |
| --- | --- | --- | --- |
| **Mental and physical health impact of the pandemic** |  |  |  |
| Increase in anxiety or stress or feelings of sadness | 74 (91.4) | 65 (86.7) | 1.05 (0.94-1.18) |
| Increase in sleep problems or poor sleep quality | 43 (53.1) | 49 (65.3) | 0.81 (0.62-1.06) |
| Increase in health problems unrelated to COVID-19 infection | 23 (28.4) | 29 (38.7) | 0.73 (0.47-1.15) |
| Decrease in physical activity or exercise | 53 (65.4) | 51 (68.9) | 0.95 (0.76-1.18) |
| Overeating or eating more unhealthy foods | 27 (33.3) | 33 (44.6) | 0.75 (0.50-1.11) |
| **Healthcare and ART access during the pandemic** |  |  |  |
| Currently on ART | 80 (98.8) | 72 (96.0) | 1.03 (0.98-1.08) |
| Difficulty accessing ART refills or pick-up | 13 (16.1) | 24 (32.4) | **0.50 (0.27-0.90)** |
| Difficulty accessing routine HIV care<sup>b</sup> | 23 (30.0) | 35 (49.3) | **0.61 (0.40-0.92)** |
| **COVID-19 Infection history** |  |  |  |
| Currently or previously symptomatic but was not tested for COVID-19 infection | 8 (9.9) | 5 (6.7) | 1.48 (0.51-4.33) |
| Tested positive for COVID-19 infection<sup>c</sup> | 2 (3.5) | 2 (4.1) | 0.86 (0.13-5.88) |
| Hospital stay due to COVID-19 infection | 0 (0) | 0 (0) | N/A |
| **Socioeconomic impact of the pandemic** |  |  |  |
| *Restricted to those who were employed prior to the pandemic (n=112, 71.8%)* |  |  |  |
| Became newly unemployed | 30 (58.8) | 21 (34.4) | **1.71 (1.13-2.59)** |
| Changed jobs | 7 (14.0) | 8 (13.1) | 1.07 (0.42-2.74) |
| Workload or work responsibilities increased | 15 (30.0) | 36 (59.0) | **0.51 (0.32-0.82)** |
| Work exposure to the general public during the pandemic | 23 (46.0) | 21 (34.4) | 1.34 (0.84-2.11) |

Abbreviations: ART = antiretroviral therapy; PWH = persons with HIV.
Prevalence ratios associated with p values < 0.05 appear in boldface
a Reference group = above secondary school
b Percentages calculated of 148 participants who attempted to access routine HIV clinical care during the pandemic; n=77, secondary school or less; n=71, above secondary school.
c Data available for n=106; n=57, secondary school or less; n=49, above secondary school.
Data was not collected on the number of participants who had a COVID-19 test.

### Knowledge of COVID-19 Infection

The majority of all participants correctly answered that the most efficacious prevention methods against COVID-19 infection were wearing a facial mask (96%), frequent handwashing (99%), avoiding close contact with people who are sick (96%) and avoiding touching the face (97%). A significantly higher proportion of participants with lower educational levels (32.1%) believed that taking antibiotics would prevent COVID-19 transmission (p=0.003). Most of the cohort correctly identified signs or symptoms related to COVID-19 infection, including cough (89.7%), fever (98.7%), shortness of breath (99.4%); data not shown.

### Socioeconomic Impact of COVID-19

Around 70% of the participants were employed prior to the pandemic. Around 46% reported being laid off from their jobs during the pandemic and nearly 40% did not follow the recommendations for telework and continued to work in close contact with the public (Table 2). Nearly one-half (51/112) reported an increase in their workload or work responsibilities. Two-thirds of those with lower educational levels became newly unemployed during the pandemic, significantly more than those with higher educational levels (58.8% vs 34.4% p=0.010; Table 2). Those with higher educational levels had significantly greater workload or work responsibilities during the pandemic compared with lower education levels (59% vs. 30%, p=0.002; Table 2). In unadjusted regression analyses, female sex was a risk factor for job loss during the pandemic (PR 1.85 [95%CI 1.27-2.69], as were lower educational levels (PR 1.62 [95%CI 1.06-2.48]), but this was not statistically significant in adjusted models. Having informal employment was an independent risk factor for unemployment during the pandemic (PR 1.58 [95%CI 1.06-2.36]), but not in the adjusted analyses (Table 3).

**Table 3:** Generalized linear models (unadjusted) for risk factors for new unemployment, mental and physical health changes during the pandemic (N=156)

|  | **Newly unemployed during pandemic<sup>a</sup>**<br>**Yes, n=51 (45.5%)** | **New or worse mental health symptoms<sup>b</sup>**<br>**Yes, n=139 (89.1%)** | **Negative physical health symptoms**<br>**Yes, n=52 (33.3%)** |
| --- | --- | --- | --- |
| **Characteristic** | **Unadjusted Prevalence Ratio (95% CI)** | **Unadjusted Prevalence Ratio (95% CI)** | **Unadjusted Prevalence Ratio (95% CI)** |
| ≥Age 50 years | 1.25<br>(0.82-1.90) | **0.89**<br>**(0.80-0.99)\*** | 1.43<br>(0.88-2.32) |
| Female sex | **1.85**<br>**(1.27-2.69)‡** | 1.09<br>(0.98-1.22) | 1.05<br>(0.67-1.65) |
| Educational level, Secondary school or less<sup>e</sup> | **1.62**<br>**(1.06-2.48)\*** | 1.05<br>(0.94-1.18) | 0.73<br>(0.47-1.15) |
| Profession type, Non-professional | 1.41<br>(0.89-2.22) | 0.89<br>(0.78-1.00) | 0.88<br>(0.50-1.54) |
| Occupation Type, Informal worker | **1.58**<br>**(1.06-2.36)\*** | 0.94<br>(0.81-1.08) | 0.62<br>(0.32-1.20) |
| Newly unemployed during the pandemic | N/A | **1.16**<br>**(1.03-1.31)\*** | 1.74<br>(0.94-3.25) |

\*p<0.05; ‡p<0.001
a Employed prior to the pandemic, n=112 (71.8%)
b New anxiety or stress or feelings of sadness since March 15, 2020

### Mental Health Impact of the COVID-19 pandemic

A majority of patients in our study expressed an increase in anxiety (64%) and stress (77%) since the beginning of the pandemic. Older age (≥50 years) and being newly unemployed during the pandemic significantly increased risk of new mental health problems in univariate analyses (PR 0.89 [CI 0.80-0.99] and PR 1.16 [1.03-1.31], respectively) but not in multivariable analyses (Table 3). There were no other significant trends related to increases in mental health symptoms. Lower education levels and employment type did not significantly increase risk of new or worse mental health symptoms during the pandemic (p>0.05 for both). Of those who had new or worse mental health symptoms during the pandemic (n=139), 60 (43.3%) sought mental health treatment or therapy, and 43.3% had difficulty accessing mental health care during the height of the pandemic with no difference by educational level (p=0.113) (Table 2).

### Physical health impact of the COVID-19 pandemic

Overall, one-third of the cohort reported an increase in health problems unrelated to COVID since the beginning of the pandemic, with no significant differences by educational level (p=0.174; Table 2). A majority of the cohort reported decreased physical activity or exercise (66.7%) with no significant differences between men and women (p=0.818 and p=0.561, respectively) and no differences by educational level (Table 2). Older age was not a significant predictor of decreased physical activity (p=0.060). Nearly 40% reported an increase in overeating or eating unhealthy foods with more men (45.6%) compared with women (28.1%) reporting this (p=0.05). Educational level and employment type were not associated with unhealthy eating habits (p>0.05).

### HIV care during the COVID-19 pandemic

Nearly all participants (97%) reported continuing to regularly take their ART medication, despite nearly one-quarter of the cohort (24%) reporting difficulty picking up their ART (Table 2). Of those who either were no longer taking their ART or had difficulty accessing ART refills (n=37), the most common reason was because of cancelled clinic appointments (26%), difficulty communicating with the HIV clinic (21%) or a lack of transportation to pick-up medications (21%). More than one-third (37.2%) reported having difficulty accessing their routine HIV medical care (Table 2), and the most common reason was that their primary HIV clinic was temporarily closed during the pandemic (46.6% [27/58]; data not shown).

## DISCUSSION

Peru has had one of the highest incidences of COVID-19 despite its early nationwide quarantine. Most ‘non-essential’ activities were suspended throughout the duration of the quarantine for nearly four months, but Peru still remains in a state of emergency with slow reactivation of the economy as of April 2021 despite confronting a second wave of cases.^16,17^ The closure of many outpatient clinics has meant that persons with chronic medical conditions requiring regular clinical care and access to life-long treatment, such as PWH, may be at increased risk for negative outcomes if their treatment is interrupted. Our study showed that most of the participants were knowledgeable in identifying correct COVID-19 infection prevention methods and COVID-19-related symptoms. We also found that the pandemic has had a large socioeconomic impact among PWH reporting significant job loss, physical and mental health burden and difficulty accessing routine HIV care or ART during the height of the pandemic.

### Knowledge and behaviors related to COVID-19

As prior studies have shown, there are discrepancies in the knowledge of infection transmission and behaviors related to COVID-19 by race, ethnicity, age and sex.^18^ One study of 225 Peruvians in the general population found that more than 90% could correctly identify COVID-19 symptoms, but more than half could not identify appropriate protective measures.^19,20^ Notably, COVID-19 knowledge increased with educational level and was associated with occupation type, independently^19^. These findings of the general population in Peru are similar to those of Peruvian PWH in our study in which the large majority could correctly identify COVID-19 symptoms highlighting the efficacy of public health messaging throughout the country.

### Mental health considerations and socioeconomic impact

The mandatory quarantines may lead to increased feelings of isolation in a group of persons who may already be vulnerable to poor mental health outcomes. In two studies of the general US poupulation,^21,22^ 40% of adults reported struggling with a mental health concern or substance use since the onset of the pandemic. Anxiety symptoms during the pandemic were three times higher (25.5% vs. 8.1%) and the prevalence of depressive symptoms four times that reported in the second quarter of 2019 (24.3% versus 6.5%).^21^

Socioeconomic challenges due to the pandemic, such as job loss or increased workload, may also create new mental health challenges or exacerbate pre-existing ones. One survey of 1,200 Peruvians in the general population published in February 2021 found that 36.6% had lost their employment since the beginning of the pandemic^23^, and in another study, 3 out of 10 people lost their jobs in Peru.^24^ Moreover, another study found that 70% of Peruvians were unable to transition work to home.^25^ Among PWH in our study, 46% reported new unemployment from March to August 2020, greater than in the general population reported to-date. However, one international study^12^ with a majority of participants from LAC (17% of the cohort were PWH) found that 11% of all participants lost their employment as a result of changes due to the pandemic, and of those who became newly unemployed, 50% had depression and 48% anxiety. Notably, men living with HIV were more likely to report expected income reductions during the pandemic compared to those without HIV (46% vs 36%; p=0.01).^12^ This highlights that PWH in Peru may be at greater risk of unemployment compared with the general population for factors including informal employment type or lower educational level.

PWH are known to be at higher risk of experiencing social isolation due to stigma and fear of rejection, particularly among older PWH.^26–28^ Among Peruvian PWH, an elevated prevalence of depression (48%-68%) has been reported,^9,29^ and more than 50% of Peruvian PWH have perceived some form of stigma or rejection associated with social isolation.^30^ One of the most striking findings from our study was that nearly half of those who were employed prior to the pandemic (46%) reported new unemployment, and significantly more with lower educational levels or having informal work reported becoming newly unemployed. This demonstrates the negative impact that quarantining measures are having on the informally employed population (nearly 70% of the Peruvian workforce).^7^

In the general population of Peru, 1 in 3 Peruvians reports anxiety or depression symptoms during the pandemic,^31^ and 144,000 Peruvians accessed mental health services from the beginning of the pandemic until July 2020^32^. Access to mental health services is crucial, particularly for PWH, during this period, however the implementation of mental health services has been challenging.^33^ We found that nearly 40% of our study participants attempted to access these services at some point during the pandemic; of them, about half had access difficulties. Some non-governmental, such as the AIDS Healthcare Foundation,^34^ have set up access to online telehealth visits with a mental health provider.^35^ Increasing the capacity of and access to such mental health services during the pandemic is crucial to help mitigate the psychological effects of the pandemic.

### Challenges to continuity of HIV care during the pandemic

In Peru, among the estimated 72,000 PWH, 64% had been diagnosed, 46% were on ART, and 36% had achieved viral suppression.^36^ However, the challenges of maintaining or improving these figures could worsen during the COVID-19 pandemic due to the re-allocation of health resources previously directed toward funding HIV care toward the inpatient care of patients with COVID-19. HIV medications are supplied and provided for free to all PWH by the Peruvian Ministry of Health, now with a three-month supply being provided to patients to help prevent shortages with ART home delivery in many regions since the beginning of the pandemic.^37^ In order to ensure medical care of PWH during a quarantine, sufficient and steady ART supplies at health care centers must be available, to achieving optimal ART adherence.^38^ However, while longer ART refills would help ensure continuous ART adherence, it also reduces the opportunity for clinical interactions with an HIV health care provider, including medication adherence counseling and viral load testing.

In one large HIV clinic in Lima, Peru, at the end of the study period (August 2020), about 50-60% of PWH were attending visits with their HIV primary care physician once every 3 months via telemedicine consultation, however many still experienced difficulties accessing telemedicine. PWH from other LAC have had similar difficulties. Santos et al. demonstrated that 23% of all PWH had difficulty accessing their HIV care provider and 18% indicated a difficulty refilling their medications.^12^ Telemedicine implementation would be optimal for continuity of care of PWH during the quarantine facilitating ART initiation and monitoring; however, barriers to implementation throughout much of Peru, including lack of consistent access to internet services and lack of necessary equipment for telemedicine within clinics, make this reality difficult to achieve.^39^ Strategizing ways ensure a steady ART supply and ART delivery to patients, and stable access to clinical care during the pandemic is fundamental to avoid an increase in HIV multimorbidity, overburdening hospitals and subsequently increasing morbidity of the general population.

Our study has limitations. First, this study was a cross-sectional telephone survey and did not have an HIV-negative control group for comparison. However, national surveys of the general Peruvian population allowed us to make comparisons to the findings of PWH in our study.^23,25,32^ Second, this was a convenience sample of middle-aged to older PWH attending two large multidisciplinary HIV clinics in an urban capital city, thus, may not be representative of the all Peruvian PWH. Third, our sample was largely educated with the majority of participants having completed secondary school, exceeding the proportion of Peruvians with at least some secondary schooling (64% female, 75.4% male),^40^. Next, most women were recruited from one of the study sites, but despite these recruitment differences, PWH did not differ greatly between study sites. On the other hand, although there have been some surveys exploring knowledge and socioeconomic, physical and mental health impacts of the pandemic in general population,^23,25^ this is the first one exploring these areas in PWH in Peru.

Our data has shown that the COVID-19 pandemic and measures taken to reduce its transmission have posed a significant mental health and socioeconomic burden on Peruvian PWH and may pose a considerable burden upon PWH in other low-to-middle income countries. As Peru enacted policies to slow the spread of COVID-19 through quarantines and shut-down of most non-essential establishments early on in the beginning of the pandemic^4^, these measures may inadvertently have negative consequences on vulnerable populations including those with HIV. These early struggles of HIV clinics in providing continued healthcare to their patients have led some clinics to develop a systematic approach to mail delivery of ART and provision of telemedicine consults, however, this is not yet standardized across much of Peru. Further efforts are needed to ensure that PWH on ART can obtain routine HIV care without increased COVID exposure by ramping up access to telehealth and ART home delivery systems. It is also crucial for those newly diagnosed with HIV during the pandemic to be linked to HIV care and treatment in a timely manner. Finally, the increase in perceived anxiety, stress and depressive symptoms observed among older PWH in our study underscores the importance of strengthening mental health resources and access via telemedicine for this vulnerable population.

## Data Availability

Data may be requested from the corresponding author.

## Acknowledgements

The authors would like to thank the HIV clinics from which participant recruitment and enrollment took place.

